# Severe cirrhosis is associated with increased surgical mortality and morbidities in patients with hip fractures: a propensity-score matched analysis using a large inpatient database

**DOI:** 10.1101/2023.04.15.23288594

**Authors:** Osamu Hamada, Jung-ho Shin, Takahiko Tsutsumi, Ayako Tsunemitsu, Noriko Sasaki, Susumu Kunisawa, Kiyohide Fushimi, Yuichi Imanaka

## Abstract

**Objective:** The aim of this study was to compare postoperative mortality and morbidities in patients with hip fractures undergoing surgery by Child-Pugh classes.

**Summary Background Data:** Advanced cirrhosis is associated with increased mortality in several types of surgery, but the impact of severity of cirrhosis on postoperative outcomes in patients with hip fractures remain unclear.

**Methods:** We analyzed data of patients with hip fracture within a large inpatient database. We performed three sets of 1:1 propensity-score matching for four groups: cases without cirrhosis, and Child-Pugh classes A, B and C. We compared in-hospital mortality, length of stay, hospitalization fee, rate of 30-day readmission and complications in the following three pairs: cases without cirrhosis vs Child-Pugh class A, Child-Pugh class A vs B, and Child-Pugh class B vs C.

**Results:** Among 833,648 eligible cases, propensity-score matching created 1,065 pairs between cases without cirrhosis vs Child-Pugh class A, 1,012 pairs between Child-Pugh class A vs B, and 489 pairs between Child-Pugh class B vs C. In-hospital mortality did not differ between cases with cirrhosis and those with Child-Pugh A classification (1.69% vs 1.41%; RD -0.28%; 95% CI: -1.34%–0.78%). In-hospital mortality was significantly higher in patients with Child-Pugh B classification than in those with A classification (1.48% vs 5.93%; RD 4.45%; 95% CI: 2.79%–6.10%), and in those with Child-Pugh C classification compared with those with B classification (6.34% vs 28.43%; RD 22.09%; 95% CI: 17.54%–26.63%). Among cases with cirrhosis, those in more severe Child-Pugh classes had longer length of stay, higher hospitalization fees and higher ratio of complications, such as acute liver failure, upper gastrointestinal bleeding and sepsis.

**Conclusions:** Our results could help to identify patients at high-risk of postoperative mortality and morbidity among those with both hip fracture and cirrhosis. Patients with Child-Pugh C classification may benefit from discussions about nonsurgical management, goals of care, and overall prognosis.

**Mini-abstract:** We conducted propensity-score matched analysis to examine mortality of groups of patients with hip fractures in a national patient database according to degree of severity of cirrhosis. Patients with higher Child-Pugh class of cirrhosis was shown to be associated with higher mortality.

## Introduction

The heavy burden of cirrhosis was shown in recent global epidemiological data.^1^ Patients with cirrhosis are living longer with more advanced disease because of improved medical and surgical management.^2^ As a result, they are at risk for hip fractures as well as other diseases and morbidities that were not previously a problem for patients with cirrhosis.^3^ Despite advances in modern surgical and intensive care treatment, perioperative mortality remains high in patients with cirrhosis that undergo surgery.^4^ Prognostic scoring systems for the underlying surgical disease in combination with the state of liver disease have therefore been investigated.^5^ Scoring systems for cirrhosis, such as Child-Pugh classification and model for end-stage liver disease (MELD), have been correlated with outcome.^6-8^ A recent review summarized 87 studies that used Child-Pugh classification or MELD. It suggested the safety of surgery for patients with Child-Pugh A classification, which had mortality <5–10% in patients undergoing non-hepatic abdominal surgery compared with 10–40% for Child-Pugh B classification and 20–100% for Child-Pugh C classification.^9^ In another study, cardiac surgery was reportedly appropriate for patients with Child-Pugh A classification and selected patients with Child-Pugh B classification, although there was a 67% risk of in-hospital mortality for patients with Child-Pugh C classification.^10^

The number of patients with hip fractures undergoing surgery increases along with a large increase in the elderly population worldwide, because the rate of hip fracture increases exponentially with age.^11^ Hip fractures greatly reduce a patient’s quality of life and physical function, and they are associated with serious mortality in the first year after injury.^12-15^ A recent systematic review combining worldwide hip fracture-related mortality rates reported one-year mortality at approximately 20%.^16^ Another single-center study showed that patients with hip fracture who were treated nonoperatively had four times higher one-year mortality after fracture than the group that underwent surgery.^17^ Surgery for hip fractures is therefore considered to be an important treatment. Most reported hip fractures occur in people older than 65 years.^18^ The global population is aging, so the number of elderly patients with cirrhosis is reported to be increasing.^19^ This combination suggests the population of patients with concurrent hip fracture and cirrhosis is increasing.^20^

Tseng et al. evaluated postoperative outcomes in 347,363 patients with hip fracture, including 2,071 with cirrhosis, from the US Nationwide Inpatient Sample. Patients with cirrhosis had higher rate of in-hospital mortality than patients without cirrhosis.^21^ Chang et al. analyzed data of 117,129 patients with hip fracture, including 4,048 with cirrhosis, from the Taiwan National Health Insurance database; they had 2–3 times higher mortality rates at three years after surgery than patients without cirrhosis.^22^ Elsewhere, Montomoli et al. analyzed data of 152,180 patients with hip fractures including 1,866 with cirrhosis from Danish registries; patients with cirrhosis had higher 30-day and 1-year mortality than patients without cirrhosis. Excess mortality was said to be due to complications of the fracture event rather than to preexisting comorbidity.^23^

To our knowledge, however, only one study has focused on the severity of cirrhosis on the mortality related to hip fractures. Hundersmarck et al. analyzed 99 patients with hip fracture with cirrhosis stratified based on MELD scores subgroup and stratified based on compensation or decompensation status, both measures of severity of cirrhosis.^24^ One-year mortality was 55% for MELD scores of 20–40 and 53% for patients with decompensated disease, compared with 16% for patients with MELD scores of 6–9 and 15% for patients with compensated disease. All in-hospital deaths were related to liver failure.^24^ However, this study is limited because of the small sample sizes.

The association between severity of cirrhosis and postoperative mortality and morbidities in patients with hip fractures requires elucidation. Understanding the impact of severity of cirrhosis for patients with hip fracture on short-term outcomes is important in determination of the surgical or non-surgical management. This real-world observational study uses a nationwide inpatient database. It aims to elucidate the impact of severity of cirrhosis in patients with hip fracture regarding postoperative outcomes.

## Methods

### Data source

We used diagnosis procedure combination (DPC) data from the database of the DPC Study Group funded by the Ministry of Health, Labour and Welfare (MHLW), Japan. The DPC/per-diem payment system (PDPS) is a Japanese payment system for inpatients in the acute phase of illness and provides survey data to the MHLW for the purpose of standardizing and improving the quality of medical care for hospitalized patients.^25^ The database contains DPC data from approximately 1,200 hospitals nationwide in 2016. The DPC data consists of discharge summaries, which include the International Classification of Diseases 10th Revision (ICD-10) Codes classifying main diagnosis, the trigger diagnosis, the most and second-most medical-resource-intensive diagnoses, comorbidities, complications during hospitalization, and various disease-specific severity classifications.^26^ The database thus includes the Child-Pugh classification for cirrhosis as well as patient details such as age, sex, body mass index (BMI), claims data of medical procedures, and daily records of drug administration.

### Study population

Included in this study were patients who met all of the following criteria: (1) admitted with hip fractures (ICD-10 Codes of S72.0–S72.2) except for open-fractures (ICD-10 Codes of S72.01, S72.11, and S72.21), (2) underwent surgery for hip fractures, (3) had complete DPC files related to medical services and medications, (4) admitted and discharged between July 1, 2010 and March 31, 2021. We excluded patients if any of the variables of the Child-Pugh classification registered as 0, which was defined as cases in which classification was not possible. Patients were classified into four groups based on the Child-Pugh classification: cases without registered Child-Pugh classification, which were regarded as cases without cirrhosis in our study, and those with Child-Pugh classes A, B and C.

### Variables

In this study, patients were categorized using the Child-Pugh classification. The Child-Pugh classification incorporates five variables which include the serum albumin, bilirubin, prothrombin time, ascites and encephalopathy. Each variable is scored one to three, with three indicating the most severe status. If one of the diagnoses registered to the DPC data include predefined diagnoses related to cirrhosis (ICD-10 Codes of I81, I82.0, I85.0, I85.9, I86.4, I98.2, I98.3, K71.7, K72.1, K72.9, K74.0-74.6, K76.5, and K76.6), registration of the Child-Pugh classification is mandatory. BMI was classified into six categories (< 18.5, 18.5-22.4, 22.5-24.9, 25.0-29.9, ≥ 30.0 kg/m^2^ and missing). Elixhauser comorbidities were coded using Quan’s protocol.^27, 28^

### Outcomes

The primary outcome of interest was in-hospital mortality. Secondary outcomes of interest were length of stay, postoperative length of stay, hospitalization fee, ratio of 30-day readmission and relevant postoperative complications, which included surgical-site infection (SSI), blood transfusion, acute liver failure, upper gastrointestinal bleeding (UGIB), respiratory failure, venous thromboembolism (VTE), acute coronary syndrome (ACS), heart failure (HF), acute renal failure, urinary tract infection (UTI), and sepsis (Supplementary Table 1).^29-31^

### Statistical analyses

We analyzed the database using three sets of propensity-score matching to address potential selection bias for four groups: cases without cirrhosis and cases with Child-Pugh A, B and C classifications. We compared the following three pairs: cases without cirrhosis vs Child-Pugh A classification, Child-Pugh A classification vs B classification, and Child-Pugh B classification vs C classification.

A one-to-one nearest neighbor propensity-score matched analysis without replacement procedure was performed using the MatchIt package^32^ in R version 4.2.1 (R Foundation for Statistical Computing, Vienna, Austria).^33^ We calculated three sets of propensity scores which were the likelihoods of being classified as Child-Pugh class A, B or C respectively in each pair based on three multivariable logistic regression models including patients’ demographic and clinical characteristics: age, sex, six BMI categories, 30 Elixhauser comorbidities, anticoagulant use and antiplatelet use. These variables were used based on previous studies and expert opinions to predict the severity of cirrhosis.^34-37^

We formed three matched pairs such that the differences in propensity scores between matched cases differ by at most a fixed distance known as a caliper width. The caliper width was set as 0.2 times of standard deviation of the logit of the propensity scores, as recommended by Austin.^35^ Balance after matching was evaluated by the absolute standardized mean difference.^36^ An absolute standardized mean difference ≤ 0.1 was considered a negligible difference and was indicative of well-balanced matching.^37^

We presented the risk difference (RD) and risk ratio (RR) as estimates of outcomes for each matched pair. Standard errors (SEs) of RD and RR, which were clustered robust SEs, were estimated using generalized linear models with log link functions.^38^ All statistical analyses were performed with R version 4.2.1 (R Foundation for Statistical Computing, Vienna, Austria).^33^

### Ethical consideration

This study was approved by the Ethics Committee, Graduate School of Medicine, Kyoto University (approval number: R0135), and was conducted in accordance with the Ethical Guidelines for Medical and Health Research Involving Human Subjects of the MHLW, Japan. According to these guidelines, the need for written informed consent was waived for this research because it did not utilize human biological specimens and all information used in the research has been anonymized.

## Results

### Study population

Included in this study were 834,262 cases of patients that underwent surgery for hip fractures during the study period. Of these, we excluded cases with any one of variables of Child-Pugh classification registered as 0 (n=614). We then identified 833,648 eligible cases (Figure 1).

**Figure 1.**
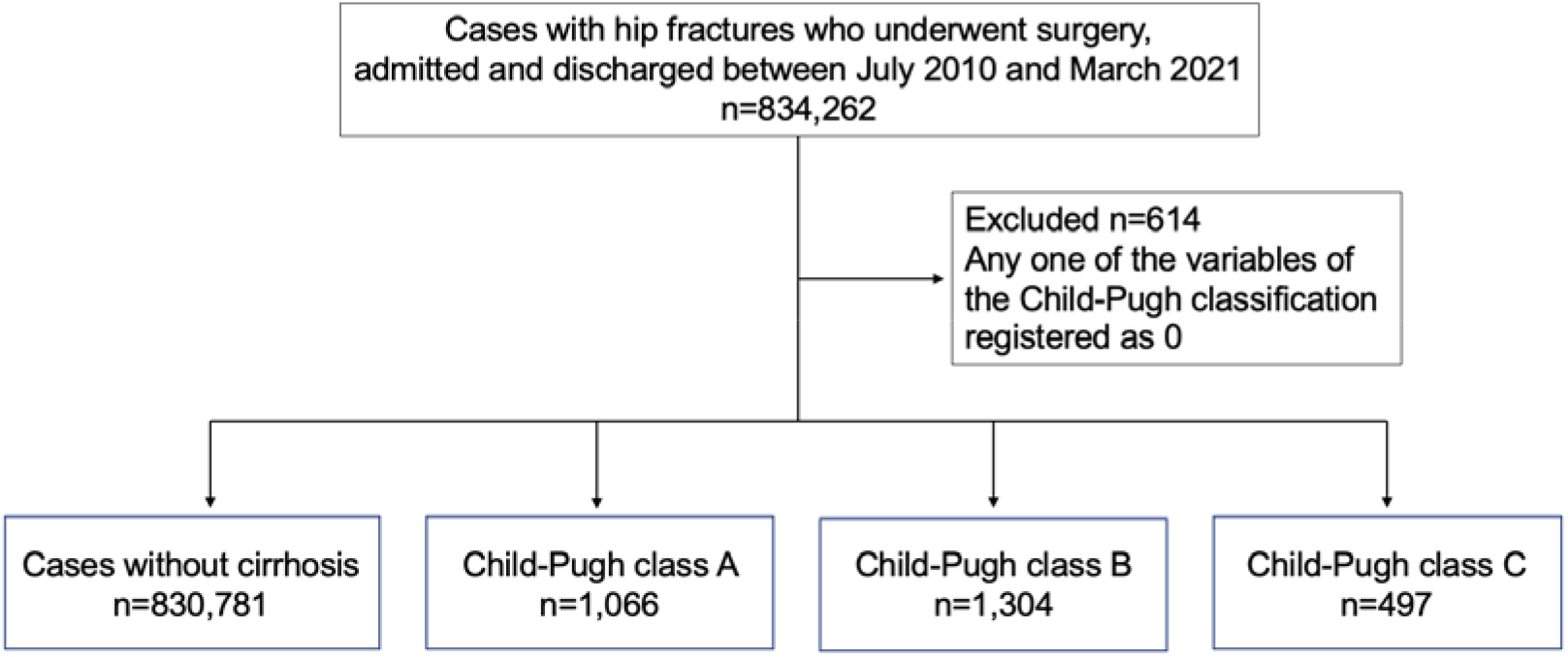
Study flow chart.

### Patient characteristics and outcomes before propensity-score matching

Before matching, the cases without cirrhosis comprised 830,781 cases, patients with Child-Pugh A classification comprised 1,066 cases, patients with Child-Pugh B classification comprised 1,304 cases, and patients with Child-Pugh C classification comprised 497 cases. The characteristics and outcomes of the patients before propensity-score matching analysis are shown in Tables 1 and 2.

**Table 1.** Study population before propensity-score matching.

|  | Cases without<br>cirrhosis | Child-Pugh class |  |  |
| --- | --- | --- | --- | --- |
|  |  | A | B | C |
| N | 830,781 | 1,066 | 1,304 | 497 |
| **Demographics** |  |  |  |  |
| age mean, SD | 82.34 (10.19) | 78.31 (9.92) | 78.31 (9.59) | 76.88 (9.96) |
| sex -female | 645,245 (77.67%) | 812 (76.17%) | 948 (72.70%) | 342 (68.81%) |
| BMI <18.5 | 220,518 (26.54%) | 254 (23.83%) | 275 (21.09%) | 96 (19.32%) |
| 18.5-22.4 | 336,810 (40.54%) | 427 (40.06%) | 507 (38.88%) | 185 (37.22%) |
| 22.5-24.9 | 124,259 (14.96%) | 175 (16.42%) | 261 (20.02%) | 106 (21.33%) |
| 25.0-29.9 | 74,156 (8.93%) | 116 (10.88%) | 155 (11.89%) | 69 (13.88%) |
| 30.0- | 10,510 (1.27%) | 19 (1.78%) | 34 (2.61%) | 15 (3.02%) |
| missing | 64,528 (7.77%) | 75 (7.04%) | 72 (5.52%) | 26 (5.23%) |
| Congestive heart failure | 68,830 (8.28%) | 62 (5.82%) | 89 (6.83%) | 34 (6.84%) |
| Cardiac arrhythmias | 56,110 (6.75%) | 43 (4.03%) | 86 (6.60%) | 23 (4.63%) |
| Valvular disease | 46,983 (5.66%) | 32 (3.00%) | 69 (5.29%) | 16 (3.22%) |
| Pulmonary circulation disorders | 3,903 (0.47%) | 4 (0.38%) | 9 (0.69%) | 1 (0.20%) |
| Peripheral vascular disorders | 10,870 (1.31%) | 11 (1.03%) | 13 (1.00%) | 0 (0.00%) |
| Hypertension | 299,953 (36.10%) | 414 (38.84%) | 384 (29.45%) | 88 (17.71%) |
| Paralysis | 3,643 (0.44%) | 2 (0.19%) | 0 (0.00%) | 1 (0.20%) |
| Other neurological disorders | 37,253 (4.48%) | 42 (3.94%) | 34 (2.61%) | 7 (1.41%) |
| Chronic pulmonary disease | 33,271 (4.00%) | 37 (3.47%) | 41 (3.14%) | 12 (2.41%) |
| Diabetes, uncomplicated | 123,624 (14.88%) | 219 (20.54%) | 237 (18.17%) | 93 (18.71%) |
| Diabetes, complicated | 27,918 (3.36%) | 51 (4.78%) | 97 (7.44%) | 26 (5.23%) |
| Hypothyroidism | 9,697 (1.17%) | 25 (2.35%) | 36 (2.76%) | 7 (1.41%) |
| Renal failure | 39,016 (4.70%) | 69 (6.47%) | 100 (7.67%) | 33 (6.64%) |
| Liver disease | 30,427 (3.66%) | 930 (87.24%) | 1155 (88.57%) | 435 (87.53%) |
| Peptic ulcer disease excluding bleeding | 24,991 (3.01%) | 43 (4.03%) | 53 (4.06%) | 17 (3.42%) |
| AIDS/HIV | 80 (0.01%) | 0 (0.00%) | 0 (0.00%) | 1 (0.20%) |
| Lymphoma | 2,336 (0.28%) | 5 (0.47%) | 3 (0.23%) | 2 (0.40%) |
| Metastatic cancer | 4,098 (0.49%) | 8 (0.75%) | 12 (0.92%) | 18 (3.62%) |
| Solid tumor without metastasis | 32,282 (3.89%) | 125 (11.73%) | 188 (14.42%) | 101 (20.32%) |
| Rheumatoid arthritis/collagen vascular disease | 18,600 (2.24%) | 46 (4.32%) | 33 (2.53%) | 5 (1.01%) |
| Coagulopathy | 2,551 (0.31%) | 32 (3.00%) | 65 (4.98%) | 28 (5.63%) |
| Obesity | 412 (0.05%) | 0 (0.00%) | 3 (0.23%) | 0 (0.00%) |
| Weight loss | 778 (0.09%) | 0 (0.00%) | 3 (0.23%) | 1 (0.20%) |
| Fluid and electrolyte disorders | 16,580 (2.00%) | 23 (2.16%) | 33 (2.53%) | 14 (2.82%) |
| Blood loss anemia | 8,105 (0.98%) | 11 (1.03%) | 19 (1.46%) | 6 (1.21%) |
| Deficiency anemia | 31,744 (3.82%) | 52 (4.88%) | 81 (6.21%) | 28 (5.63%) |
| Alcohol abuse | 2,587 (0.31%) | 26 (2.44%) | 52 (3.99%) | 28 (5.63%) |
| Drug abuse | 35 (0.00%) | 0 (0.00%) | 0 (0.00%) | 0 (0.00%) |
| Psychoses | 32,883 (3.96%) | 20 (1.88%) | 25 (1.92%) | 8 (1.61%) |
| Depression | 26,278 (3.16%) | 36 (3.38%) | 33 (2.53%) | 7 (1.41%) |
| Anticoagulant use | 222,278 (26.76%) | 227 (21.29%) | 304 (23.31%) | 98 (19.72%) |
| Antiplatelet use | 163,128 (19.64%) | 121 (11.35%) | 100 (7.67%) | 28 (5.63%) |

SD, standard deviations; BMI, body mass index; AIDS/HIV, acquired immune deficiency syndrome/human immunodeficiency virus.

**Table 2.** Outcomes before propensity-score matching.

|  | Cases without cirrhosis | Child-Pugh class |  |  |
| --- | --- | --- | --- | --- |
|  |  | A | B | C |
| N | 830,781 | 1,066 | 1,304 | 497 |
| **Outcome** |  |  |  |  |
| In-hospital death | 13,741 (1.65%) | 15 (1.41%) | 75 (5.75%) | 141 (28.37%) |
| Length of stay mean, SD | 35.90 (26.78) | 39.0 (27.92) | 41.9 (31.16) | 52.5 (38.89) |
| Postoperative days mean, SD | 30.92 (25.66) | 33.3 (25.71) | 35.8 (29.03) | 45.9 (37.50) |
| 30-day readmission | 28,663 (3.45%) | 59 (5.53%) | 111 (8.51%) | 51 (10.26%) |
| Hospitalization fee mean JPY, SD | 1,772,150 (863,700) | 1,943,972 (996,936) | 2,093,716 (1,096,777) | 2,528,731 (1,443,841) |
| **Complications** |  |  |  |  |
| **Surgical** |  |  |  |  |
| Surgical site infection | 423 (0.05%) | 2 (0.19%) | 1 (0.08%) | 1 (0.20%) |
| Blood transfusion | 314,300 (37.83%) | 524 (49.16%) | 841 (64.49%) | 393 (79.07%) |
| **Non-surgical** |  |  |  |  |
| Acute liver failure | 86 (0.01%) | 31 (2.91%) | 90 (6.90%) | 88 (17.71%) |
| Upper gastrointestinal bleeding | 24,506 (2.95%) | 137 (12.85%) | 153 (11.73%) | 88 (17.71%) |
| Respiratory failure | 33,831 (4.07%) | 42 (3.94%) | 57 (4.37%) | 48 (9.66%) |
| Venous thromboembolism | 31,761 (3.82%) | 43 (4.03%) | 45 (3.45%) | 17 (3.42%) |
| Acute coronary syndrome | 10,759 (1.30%) | 13 (1.22%) | 9 (0.69%) | 3 (0.60%) |
| Heart failure | 11,454 (1.38%) | 15 (1.41%) | 29 (2.22%) | 7 (1.41%) |
| Stroke | 6,076 (0.73%) | 5 (0.47%) | 5 (0.38%) | 0 (0.00%) |
| Acute renal failure | 1,402 (0.17%) | 3 (0.28%) | 7 (0.54%) | 11 (2.21%) |
| Urinary tract infection | 28,943 (3.48%) | 43 (4.03%) | 50 (3.83%) | 20 (4.02%) |
| Sepsis | 11,294 (1.36%) | 28 (2.63%) | 35 (2.68%) | 34 (6.84%) |

SD, standard deviations: JPY, Japanese yen.

Our study population consisted of cases undergoing surgery, so mean ages (standard deviation) tended to be younger in more severe Child-Pugh classes. Patients with Child-Pugh A classification had a mean of 78.31 (9.92) years, those with Child-Pugh B classification had a mean of 78.31 (9.59) years, and those with Child-Pugh C classification had a mean of 76.88 (9.96) years. Ratio of patients with metastatic cancer tended to be higher in more severe Child-Pugh classes. Patients with Child-Pugh A classification had a rate of 0.75%, those with Child-Pugh B classification had a rate of 0.92%, and those with Child-Pugh C classification had a rate of 3.62%. The ratio of solid tumors without metastasis tended to be higher in more severe Child-Pugh classes. Patients with Child-Pugh A classification had a rate of 11.73%, those with Child-Pugh B classification had a rate of 14.42%, and those with Child-Pugh C classification had a rate of 20.32%. Ratio of coagulopathy including thrombocytopenia showed a tendency to be higher in more severe Child-Pugh classes. Patients with Child-Pugh A classification had a rate of 3.00%, those with Child-Pugh B classification had a rate of 4.98%, and those with Child-Pugh C classification had a rate of 5.63% (Table 1).

Cases in more severe Child-Pugh classes of cirrhosis tended to have higher ratio of in-hospital mortality and complications. In-hospital mortality was 1.41%, 5.75% and 28.37% in Child-Pugh A classification, Child-Pugh B classification and Child-Pugh C classification, respectively. The rates of complications such as acute liver failure were 2.91%, 6.90% and 17.71% in Child-Pugh A, B and C classes, respectively (Table 2).

### Patient characteristics and outcomes after propensity-score matching

After propensity-score matching, the number of pairs were 1,065 (cases without cirrhosis vs Child-Pugh A classification), 1,012 (Child-Pugh class A vs B), and 489 (Child-Pugh class B vs C) (Table 3). The ratio of matched cases were 99.9%, 77.6% and 98.4% respectively. The characteristics in each pair before and after the propensity-score matching are shown in Supplementary Tables 2–4. Absolute values of standardized differences for each variable were ≤ 0.1 after propensity score matching.

**Table 3.**
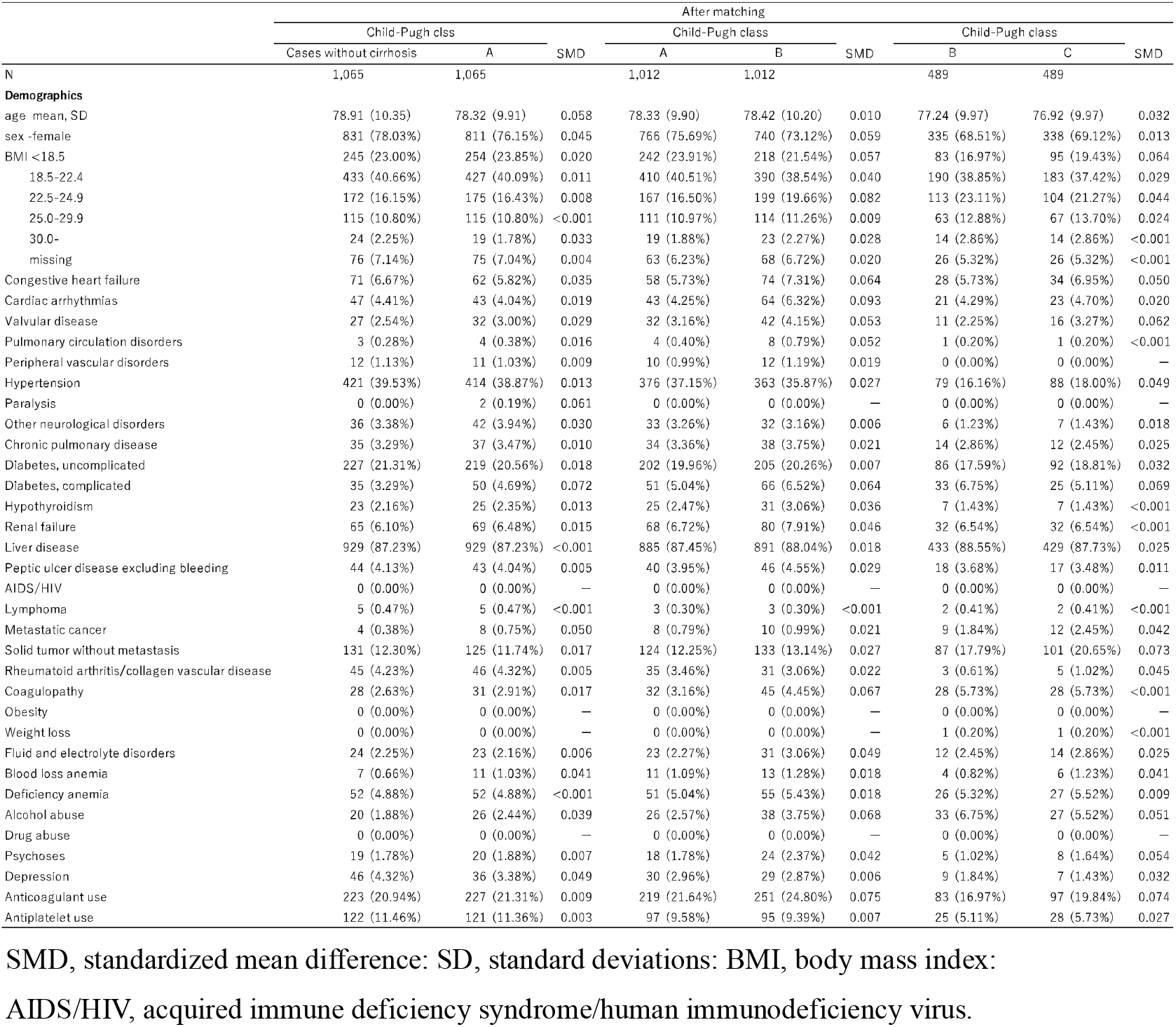
Study population after propensity-score matching.

The outcomes, and estimated RDs and RRs of matched cases are shown in Table 4 and Supplemental Tables 5–7. In-hospital mortality, which was the primary outcome, was 1.69% in cases without cirrhosis and 1.41% in Child-Pugh A classification (RD -0.28%; 95% CI: -1.34%–0.78%; RR 0.83; 95% CI: 0.42–1.65). In-hospital mortality was higher with statistical significance in patients with Child-Pugh B classification than those with Child-Pugh A classification (1.48% vs 5.93%; RD 4.45%; 95% CI: 2.79%–6.10%; RR 4.00; 95% CI: 2.27–7.05), and in those with Child-Pugh C classification compared with those with Child-Pugh B classification (6.34% vs 28.43%; RD 22.09%; 95% CI: 17.54%–26.63%; RR 4.48; 95% CI: 3.10–6.48) (Table 4 and Supplementary Tables 5–7).

**Table 4.** Outcomes after propensity-score matching.

| Outcome | Child-Pugh class A vs cases without cirrhosis |  | Child-Pugh class B vs A |  | Child-Pugh class C vs B |  |
| --- | --- | --- | --- | --- | --- | --- |
|  | RD, % (95%CI) | RR (95%CI) | RD, % (95%CI) | RR (95%CI) | RD, % (95%CI) | RR (95%CI) |
| **Outcome** |  |  |  |  |  |  |
| In-hospital death | -0.28 (-1.34–0.78) | 0.83 (0.42–1.65) | **4.45 (2.79–6.10)** | **4.00 (2.27–7.05)** | **22.09 (17.54–26.63)** | **4.48 (3.10–6.48)** |
| Length of stay mean | -0.12 (-2.56–2.33) | 1.00 (0.94–1.06) | **3.77 (1.09–6.45)** | **1.10 (1.03–1.17)** | **12.46 (8.41–16.52)** | **1.31 (1.20–1.43)** |
| Postoperative days mean | -0.86 (-3.17–1.44) | 0.97 (0.91–1.04) | **3.31 (0.84–5.77)** | **1.10 (1.02–1.18)** | **11.89 (7.94–15.84)** | **1.35 (1.22–1.48)** |
| 30-day readmission | -1.13 (-3.14–0.89) | 0.83 (0.60–1.16) | **2.37 (0.18–4.56)** | **1.41 (1.02–1.95)** | 1.23 (-2.49–4.95) | 1.13 (0.78–1.66) |
| Hospitalization fee mean JPY | **83,471 (4,155–162,788)** | **1.04 (1.00–1.09)** | **164,730 (73,062–256,399)** | **1.08 (1.04–1.14)** | **451,560 (292,234–610,886)** | **1.22 (1.14–1.30)** |
| **Complications** |  |  |  |  |  |  |
| **Surgical** |  |  |  |  |  |  |
| Surgical site infection | 0.19 (-0.07–0.45) | — — | -0.10 (-0.43–0.24) | 0.50 (0.05–5.52) | 0.00 (-0.57–0.57) | 1.00 (0.06–16.03) |
| Blood transfusion | **9.86 (5.76–13.95)** | **1.25 (1.14–1.37)** | **14.82 (10.54–19.10)** | **1.30 (1.20–1.41)** | **15.34 (9.67–21.00)** | **1.24 (1.14–1.35)** |
| **Non-surgical** |  |  |  |  |  |  |
| Acute liver failure | **2.91 (1.90–3.92)** | — — | **4.15 (2.27–6.03)** | **2.45 (1.60–3.74)** | **9.00 (4.71–13.28)** | **2.00 (1.42–2.82)** |
| Upper gastrointestinal bleeding | **9.95 (7.67–12.23)** | **4.42 (3.01–6.50)** | -0.99 (-3.82–1.85) | 0.92 (0.74–1.16) | **6.54 (2.08–11.01)** | **1.60 (1.16–2.23)** |
| Respiratory failure | 1.22 (-0.31–2.75) | 1.45 (0.91–2.31) | 0.79 (-0.92–2.50) | 1.22 (0.80–1.86) | **5.11 (2.01–8.21)** | **2.19 (1.34–3.59)** |
| Venous thromboembolism | 0.00 (-1.63–1.63) | 1.00 (0.67–1.50) | -0.59 (-2.30–1.12) | 0.86 (0.55–1.34) | 0.00 (-2.27–2.27) | 1.00 (0.52–1.92) |
| Acute coronary syndrome | 0.38 (-0.49–1.24) | 1.44 (0.62–3.38) | -0.40 (-1.22–0.43) | 0.64 (0.25–1.64) | 0.00 (-0.98–0.98) | 1.00 (0.20–4.96) |
| Heart failure | 0.38 (-0.56–1.31) | 1.36 (0.63–2.97) | 0.69 (-0.49–1.87) | 1.47 (0.76–2.83) | 0.00 (-1.27–1.27) | 1.00 (0.29–3.46) |
| Stroke | 0.09 (-0.46–0.65) | 1.25 (0.34–4.66) | -0.00 (-0.55–0.55) | 1.00 (0.25–4.00) | — — | — — |
| Acute renal failure | 0.09 (-0.32–0.51) | 1.50 (0.25–8.98) | 0.40 (-0.15–0.94) | 3.00 (0.61–14.88) | **1.64 (0.38–2.90)** | **9.00 (1.14–71.19)** |
| Urinary tract infection | -0.19 (-1.88–1.50) | 0.96 (0.64–1.44) | -0.00 (-1.67–1.67) | 1.00 (0.64–1.56) | 0.82 (-1.52–3.16) | 1.25 (0.66–2.37) |
| Sepsis | **1.41 (0.26–2.56)** | **2.15 (1.13–4.09)** | 0.30 (-1.17–1.76) | 1.11 (0.66–1.87) | **4.09 (1.45–6.73)** | **2.43 (1.34–4.41)** |

RD, risk difference; RR, risk ratio; CI, confidence interval; SD, standard deviations; JPY, Japanese yen. Bold indicates statistically significant.

Among the secondary outcomes, length of stay and postoperative length of stay tended to be longer, and ratio of 30-day readmission and hospitalization fee tended to be higher in cases with more severe Child-Pugh classes (Table 4 and Supplementary Tables 5–7). Regarding complications, there was no statistically significant difference in ratio of SSI, VTE, ACS, HF, stroke and UTI between groups. Meanwhile, patients with Child-Pugh C classification had higher ratio of blood transfusion, acute liver failure, UGIB, respiratory failure, acute renal failure and sepsis than those of Child-Pugh B classification (Table 4 and Supplementary Table 7). Of these, rate of acute liver failure was higher with statistical significance in the patients with Child-Pugh A classification than cases without cirrhosis (0% vs 2.91%; RD 2.91%; 95% CI: 1.90%–3.92%), in the Child-Pugh B classification than those with Child-Pugh A classification (2.87% vs 7.02%; RD 4.15%; 95% CI: 2.27%–6.03%; RR 2.45; 95% CI: 1.60–3.74), and in those with Child-Pugh C classification compared with those with Child-Pugh B classification (9.00% vs 18.00%; RD 9.00%; 95% CI: 4.71%–13.28%; RR 2.00; 95% CI: 1.42–2.82) (Table 4 and Supplementary Tables 5–7).

## Discussion

We compared postoperative outcomes in patients in a nationwide inpatient database with hip fractures undergoing surgery by Child-Pugh classes representing severity of cirrhosis. Among cases with cirrhosis, cases in more severe Child-Pugh classes showed higher in-hospital mortality, longer length of overall stay and postoperative length of stay and higher hospitalization fees. Notably, significant differences were shown between Child-Pugh classes B and C. There was no statistically significant difference between the groups in ratios of SSI, VTE, ACS, HF, stroke or UTI.

Patients in more severe Child-Pugh classes of cirrhosis were associated with higher in-hospital mortality. Severity of cirrhosis was significantly associated with higher postoperative mortality in a recent retrospective cohort study where patients with hip fracture with cirrhosis were stratified based on MELD score subgroups.^24^ In this study, all in-hospital deaths were related to liver failure.^24^ Elsewhere, a relationship between the development and outcome of acute-on-chronic liver failure after surgical interventions.^42^ Acute complications, such as gastrointestinal bleeding, bacterial infections, development or worsening of ascites, encephalopathy, and nonobstructive jaundice, led to acute decompensation episodes.^43^ Acute decompensation may progress further to acute-on-chronic liver failure, which is associated with high short-term mortality.^44^ In the current study there were higher ratios in cases with Child-Pugh C classification of acute liver failure, UGIB, sepsis and respiratory failure including pneumonia especially compared with patients with Child-Pugh B classification. This may be related to higher in-hospital mortality as described in previous studies.^24,^ 42-44

Our results showed patients in more severe Child-Pugh classes had longer overall length of stay and postoperative length of stay and higher hospitalization fees than less severe classes. A recent retrospective cohort study reported that patients with decompensated cirrhosis undergoing orthopedic procedures had higher in-hospital mortality, longer lengths of stay and higher hospitalization costs.^45^ However, there was no discussion of causes of longer length of stay and higher hospitalization cost in this study.^45^ Our study nonetheless had similar findings of length of stay and hospitalization fee. This similarity may be explained by cases with more severe Child-Pugh class tending to have higher rate of complications in our study. These associations are clinically plausible, and a previous report also showed that the complications were associated with increased length of stay and hospitalization cost.^46^

Our study showed no significant difference in rates of VTE, SSI or UTI, whereas cases with Child-Pugh C classification had higher rate of respiratory failure including pneumonia. A previous study analyzing patients with hip fracture with cirrhosis stratified based on MELD scores subgroups showed that the rates of postoperative complications including symptomatic thromboembolism, SSI, pneumonia and UTI were not different among MELD score of 6-19, MELD score of 10-19 and MELD score of 20-40 groups.^24^ Our study similarly showed that the ratio of most complications was not different between groups.

Our result showed higher ratio of respiratory failure including pneumonia in cases with Child-Pugh C classification than those with Child-Pugh B classification. A recent retrospective cohort study compared patients with and without postoperative pneumonia, and liver disease was an independent risk factors for postoperative pneumonia following surgically-treated hip fracture in geriatric patients (Odds ratio 1.61; 95% CI: 1.15–5.15).^47^ Other studies reported that patients with cirrhosis had a higher incidence of postoperative pneumonia than those without.^48-50^ These associations are clinically plausible because infections were reported to be more frequent among the patients with cirrhosis as an effect of the immune system suppression related to the cirrhosis.^51, 52^ Altogether, our results are in line with those in these previous studies.^47-52^

Our analysis demonstrated that cases with more advanced cirrhosis had a tendency to have higher rates of blood transfusion. Patients with advanced cirrhosis were reported to have higher prevalence of coagulopathy and thrombocytopenia.^53^ Although information on the severity of the coagulopathy and thrombocytopenia was unavailable, we adjusted coagulopathy, thrombocytopenia, anticoagulant and antiplatelet use between groups using propensity-score. Therefore, more advanced cirrhosis may have increased bleeding tendency which could not be explained by adjusted coagulopathy, thrombocytopenia, and the use of anticoagulants and antiplatelet. Cases in more severe Child-Pugh classes showed higher ratio of UGIB, and patients with advanced cirrhosis were reported to have higher rates of hemorrhage and hematomas, even after minor invasive procedures.^54^ Our results with higher ratio of blood transfusion in cases with more advanced cirrhosis may be explained by increased bleeding tendency and higher ratio of UGIB and surgery by itself.

We found no statistically significant difference in rate of ACS, HF and strokes. A previous study that analyzed patients with hip fracture with cirrhosis stratified based on MELD scores subgroups only evaluated rates of postoperative complications including symptomatic thromboembolism, SSI, pneumonia and UTI.^24^ Our propensity-matched comparison may add details on rates of postoperative complications in patients with hip fracture with cirrhosis.

Our study had several limitations. First, this was a retrospective observational study and there may have been unmeasured covariates. However, ethical issues make it impossible to conduct a randomized controlled trial for our study population. Second, we were unable to analyze the impact of cirrhosis etiology owing to data availability. Cirrhosis etiology may account for outcomes such as in-hospital mortality and acute liver failure. In a recent study, etiology did not appear to play a major role in the development of acute-on-chronic liver failure after surgery, which was associated with high short-term mortality,^42^ although it was strongly associated with acute-on-chronic liver failure in another study.^44^ Third, the DPC database did not include information on factors such as surgeon and anesthesiologist volumes, and multidisciplinary collaboration system with surgeons and specialists in each hospital. A systematic review reported the surgeon volume-outcome relationship on 15 surgical procedures and conditions.^55^ Elsewhere, those who received care from high-volume anesthesiologists had a lower risk of adverse postoperative outcomes compared with those who received care from low-volume anesthesiologists.^56^ Some studies reported the importance of multidisciplinary management of patients with cirrhosis.^57,58^ These factors may be associated with mortality and morbidities in our study population.

Despite several limitations, this was nonetheless a real-world observational study using a nationwide inpatient database with large sample size. Our results suggest importance of the risk stratification by severity of cirrhosis to inform on perioperative optimization and facilitate risk-benefit discussions. Patients with Child-Pugh A and B classification are known to be able to undergo most surgeries with a reasonable safety profile.^53^ In fact, in-hospital mortality in Child-Pugh A and B classes in our study was similar to previously-reported in-hospital mortality in patients with hip fracture ranging from 1.52–11.4%.^59-61^ On the other hand, like other surgeries, postoperative in-hospital mortality was higher in cases with higher Child-Pugh classes, notably in Child-Pugh C classification than those of Child-Pugh B classification. A recent systematic review reported a one-year mortality rate of 22% with a range from 2.4%–34.8%.^16^ A single center study showed that patients with hip fracture who were treated nonoperatively had four times higher one-year mortality than the operative group.^17^ However, our results showed cases with Child-Pugh C classification had significantly higher in-hospital mortality than those of Child-Pugh B classification. Comprehensive shared decision making may be needed for patients with Child-Pugh C classification and should include consideration of surgical risk according to the severity of cirrhosis, risk of treatment without surgery, the patient’s frailty, and their personal wishes.

## Conclusions

This propensity score-matched analysis of patients with concurrent hip fracture and cirrhosis that underwent surgery demonstrated significant differences in risk of in-hospital mortality and morbidities according to severity of cirrhosis. Patients with Child-Pugh C classification may need optimization and discussions about nonsurgical management, goals of care, and overall prognosis.

## Data Availability

All data produced in the present study are available upon reasonable request to the authors.

---

**Supplementary Table 1.**
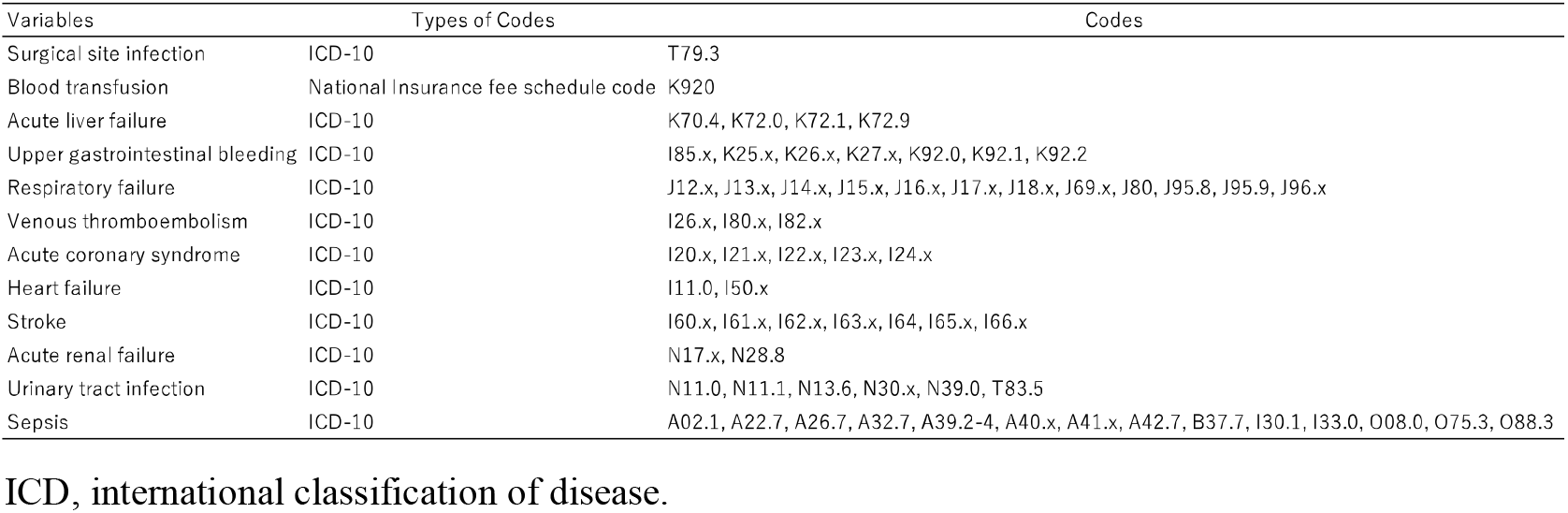
Variables, Types of Codes, and Corresponding Codes.

**Supplementary Table 2.**
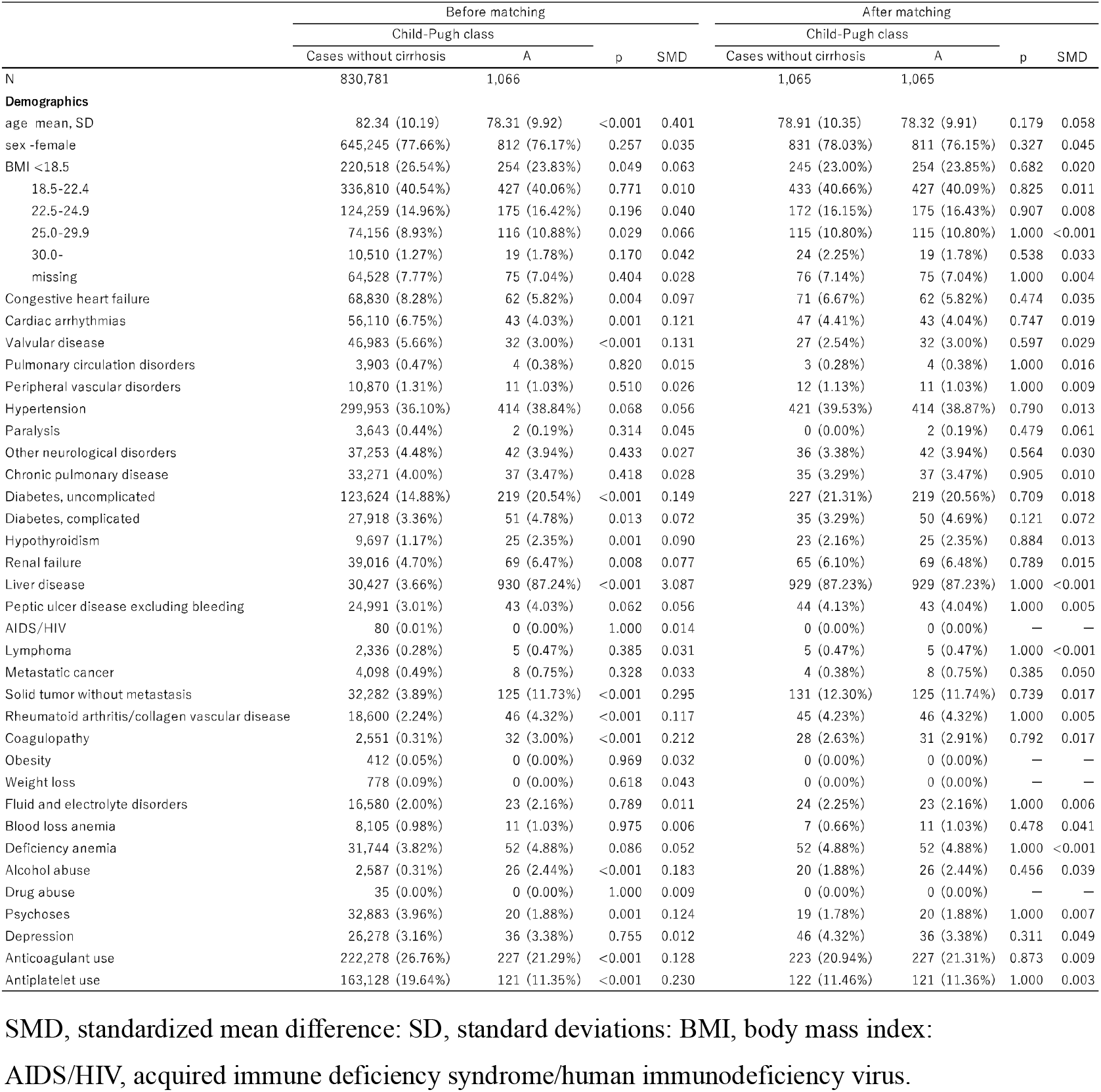
Study population before and after propensity-score matching for cases without cirrhosis and Child-Pugh class A.

**Supplementary Table 3.**
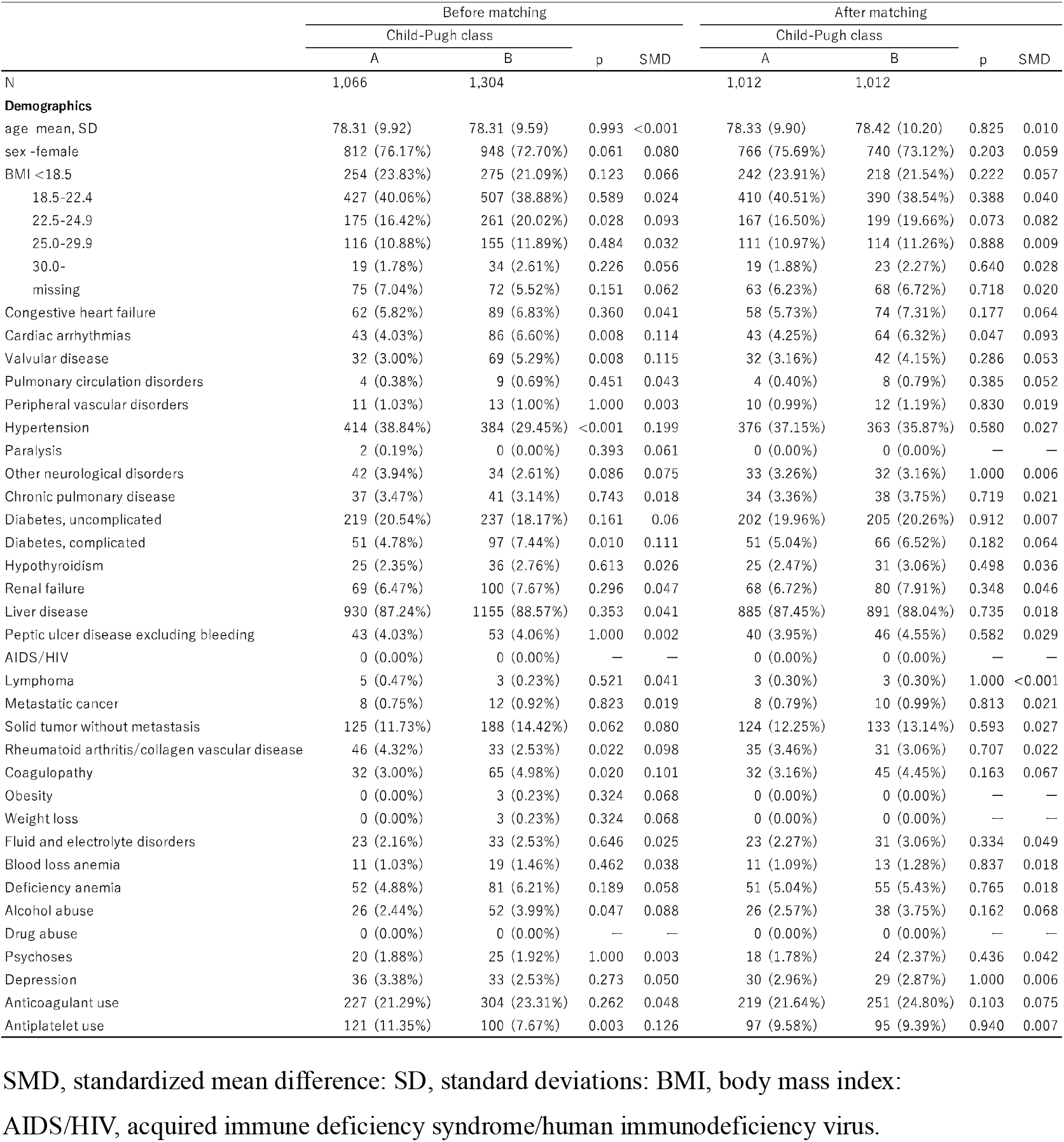
Study population before and after propensity-score matching for Child-Pugh classes A and B.

**Supplementary Table 4.** Study population before and after propensity-score matching for Child-Pugh classes B and C.

|  | Before matching |  |  |  | After matching |  |  |  |
| --- | --- | --- | --- | --- | --- | --- | --- | --- |
|  | Child-Pugh class |  | p | SMD | Child-Pugh class |  | p | SMD |
|  | B | C |  |  | B | C |  |  |
| N | 1,304 | 497 |  |  | 489 | 489 |  |  |
| **Demographics** |  |  |  |  |  |  |  |  |
| age mean, SD | 78.31 (9.59) | 76.88 (9.96) | 0.005 | 0.145 | 77.24 (9.97) | 76.92 (9.97) | 0.621 | 0.032 |
| sex -female | 948 (72.70%) | 342 (68.81%) | 0.115 | 0.086 | 335 (68.51%) | 338 (69.12%) | 0.890 | 0.013 |
| BMI <18.5 | 275 (21.09%) | 96 (19.32%) | 0.443 | 0.044 | 83 (16.97%) | 95 (19.43%) | 0.362 | 0.064 |
| 18.5-22.4 | 507 (38.88%) | 185 (37.22%) | 0.554 | 0.034 | 190 (38.85%) | 183 (37.42%) | 0.693 | 0.029 |
| 22.5-24.9 | 261 (20.02%) | 106 (21.33%) | 0.580 | 0.032 | 113 (23.11%) | 104 (21.27%) | 0.538 | 0.044 |
| 25.0-29.9 | 155 (11.89%) | 69 (13.88%) | 0.286 | 0.060 | 63 (12.88%) | 67 (13.70%) | 0.778 | 0.024 |
| 30.0- | 34 (2.61%) | 15 (3.02%) | 0.751 | 0.025 | 14 (2.86%) | 14 (2.86%) | 1.000 | <0.001 |
| missing | 72 (5.52%) | 26 (5.23%) | 0.899 | 0.013 | 26 (5.32%) | 26 (5.32%) | 1.000 | <0.001 |
| Congestive heart failure | 89 (6.83%) | 34 (6.84%) | 1.000 | 0.001 | 28 (5.73%) | 34 (6.95%) | 0.512 | 0.050 |
| Cardiac arrhythmias | 86 (6.60%) | 23 (4.63%) | 0.146 | 0.086 | 21 (4.29%) | 23 (4.70%) | 0.877 | 0.020 |
| Valvular disease | 69 (5.29%) | 16 (3.22%) | 0.084 | 0.103 | 11 (2.25%) | 16 (3.27%) | 0.435 | 0.062 |
| Pulmonary circulation disorders | 9 (0.69%) | 1 (0.20%) | 0.372 | 0.073 | 1 (0.20%) | 1 (0.20%) | 1.000 | <0.001 |
| Peripheral vascular disorders | 13 (1.00%) | 0 (0.00%) | 0.055 | 0.142 | 0 (0.00%) | 0 (0.00%) | — | — |
| Hypertension | 384 (29.45%) | 88 (17.71%) | <0.001 | 0.279 | 79 (16.16%) | 88 (18.00%) | 0.497 | 0.049 |
| Paralysis | 0 (0.00%) | 1 (0.20%) | 0.616 | 0.064 | 0 (0.00%) | 0 (0.00%) | — | — |
| Other neurological disorders | 34 (2.61%) | 7 (1.41%) | 0.178 | 0.086 | 6 (1.23%) | 7 (1.43%) | 1.000 | 0.018 |
| Chronic pulmonary disease | 41 (3.14%) | 12 (2.41%) | 0.507 | 0.044 | 14 (2.86%) | 12 (2.45%) | 0.842 | 0.025 |
| Diabetes, uncomplicated | 237 (18.17%) | 93 (18.71%) | 0.845 | 0.014 | 86 (17.59%) | 92 (18.81%) | 0.679 | 0.032 |
| Diabetes, complicated | 97 (7.44%) | 26 (5.23%) | 0.120 | 0.091 | 33 (6.75%) | 25 (5.11%) | 0.343 | 0.069 |
| Hypothyroidism | 36 (2.76%) | 7 (1.41%) | 0.132 | 0.095 | 7 (1.43%) | 7 (1.43%) | 1.000 | <0.001 |
| Renal failure | 100 (7.67%) | 33 (6.64%) | 0.519 | 0.040 | 32 (6.54%) | 32 (6.54%) | 1.000 | <0.001 |
| Liver disease | 1155 (88.57%) | 435 (87.53%) | 0.592 | 0.032 | 433 (88.55%) | 429 (87.73%) | 0.767 | 0.025 |
| Peptic ulcer disease excluding bleeding | 53 (4.06%) | 17 (3.42%) | 0.620 | 0.034 | 18 (3.68%) | 17 (3.48%) | 1.000 | 0.011 |
| AIDS/HIV | 0 (0.00%) | 1 (0.20%) | 0.616 | 0.064 | 0 (0.00%) | 0 (0.00%) | — | — |
| Lymphoma | 3 (0.23%) | 2 (0.40%) | 0.904 | 0.031 | 2 (0.41%) | 2 (0.41%) | 1.000 | <0.001 |
| Metastatic cancer | 12 (0.92%) | 18 (3.62%) | <0.001 | 0.182 | 9 (1.84%) | 12 (2.45%) | 0.659 | 0.042 |
| Solid tumor without metastasis | 188 (14.42%) | 101 (20.32%) | 0.003 | 0.156 | 87 (17.79%) | 101 (20.65%) | 0.291 | 0.073 |
| Rheumatoid arthritis/collagen vascular disease | 33 (2.53%) | 5 (1.01%) | 0.067 | 0.116 | 3 (0.61%) | 5 (1.02%) | 0.723 | 0.045 |
| Coagulopathy | 65 (4.98%) | 28 (5.63%) | 0.662 | 0.029 | 28 (5.73%) | 28 (5.73%) | 1.000 | <0.001 |
| Obesity | 3 (0.23%) | 0 (0.00%) | 0.672 | 0.068 | 0 (0.00%) | 0 (0.00%) | — | — |
| Weight loss | 3 (0.23%) | 1 (0.20%) | 1.000 | 0.006 | 1 (0.20%) | 1 (0.20%) | 1.000 | <0.001 |
| Fluid and electrolyte disorders | 33 (2.53%) | 14 (2.82%) | 0.861 | 0.018 | 12 (2.45%) | 14 (2.86%) | 0.842 | 0.025 |
| Blood loss anemia | 19 (1.46%) | 6 (1.21%) | 0.857 | 0.022 | 4 (0.82%) | 6 (1.23%) | 0.751 | 0.041 |
| Deficiency anemia | 81 (6.21%) | 28 (5.63%) | 0.727 | 0.024 | 26 (5.32%) | 27 (5.52%) | 1.000 | 0.009 |
| Alcohol abuse | 52 (3.99%) | 28 (5.63%) | 0.165 | 0.077 | 33 (6.75%) | 27 (5.52%) | 0.505 | 0.051 |
| Drug abuse | 0 (0.00%) | 0 (0.00%) | — | — | 0 (0.00%) | 0 (0.00%) | — | — |
| Psychoses | 25 (1.92%) | 8 (1.61%) | 0.812 | 0.023 | 5 (1.02%) | 8 (1.64%) | 0.577 | 0.054 |
| Depression | 33 (2.53%) | 7 (1.41%) | 0.206 | 0.081 | 9 (1.84%) | 7 (1.43%) | 0.801 | 0.032 |
| Anticoagulant use | 304 (23.31%) | 98 (19.72%) | 0.115 | 0.088 | 83 (16.97%) | 97 (19.84%) | 0.283 | 0.074 |
| Antiplatelet use | 100 (7.67%) | 28 (5.63%) | 0.162 | 0.082 | 25 (5.11%) | 28 (5.73%) | 0.778 | 0.027 |

SMD, standardized mean difference; SD, standard deviations; BMI, body mass index;
AIDS/HIV, acquired immune deficiency syndrome/human immunodeficiency virus.

**Supplementary Table 5.** Outcomes after propensity-score matching for cases without cirrhosis and Child-Pugh class A.

|  | Child-Pugh class |  | RD, % (95%CI) | P | RR (95%CI) | P |
| --- | --- | --- | --- | --- | --- | --- |
|  | Cases without cirrhosis | A |  |  |  |  |
| N | 1,065 | 1,065 |  |  |  |  |
| **Outcome** |  |  |  |  |  |  |
| In-hospital death | 18 (1.69%) | 15 (1.41%) | -0.28 (-1.34–0.78) | 0.602 | 0.83 (0.42–1.65) | 0.602 |
| Length of stay mean, SD | 39.15 (29.76) | 39.03 (27.93) | -0.12 (-2.56–2.33) | 0.924 | 1.00 (0.94–1.06) | 0.924 |
| Postoperative days mean, SD | 34.16 (28.76) | 33.29 (25.72) | -0.86 (-3.17–1.44) | 0.463 | 0.97 (0.91–1.04) | 0.462 |
| 30-day readmission | 71 (6.67%) | 59 (5.54%) | -1.13 (-3.14–0.89) | 0.273 | 0.83 (0.60–1.16) | 0.274 |
| Hospitalization fee mean JPY, SD | 1,861,070 (879,277) | 1,944,542 (997,230) | **83,471 (4,155–162,788)** | 0.039 | **1.04 (1.00–1.09)** | 0.039 |
| **Complications** |  |  |  |  |  |  |
| **Surgical** |  |  |  |  |  |  |
| Surgical site infection | 0 (0.00%) | 2 (0.19%) | 0.19 (-0.07–0.45) | 0.157 | — — | — |
| Blood transfusion | 418 (39.25%) | 523 (49.11%) | **9.86 (5.76–13.95)** | <0.001 | **1.25 (1.14–1.37)** | <0.001 |
| **Non-surgical** |  |  |  |  |  |  |
| Acute liver failure | 0 (0.00%) | 31 (2.91%) | **2.91 (1.90–3.92)** | <0.001 | — — | — |
| Upper gastrointestinal bleeding | 31 (2.91%) | 137 (12.86%) | **9.95 (7.67–12.23)** | <0.001 | **4.42 (3.01–6.50)** | <0.001 |
| Respiratory failure | 29 (2.72%) | 42 (3.94%) | 1.22 (-0.31–2.75) | 0.117 | 1.45 (0.91–2.31) | 0.120 |
| Venous thromboembolism | 43 (4.04%) | 43 (4.04%) | 0.00 (-1.63–1.63) | 1.000 | 1.00 (0.67–1.50) | 1.000 |
| Acute coronary syndrome | 9 (0.85%) | 13 (1.22%) | 0.38 (-0.49–1.24) | 0.394 | 1.44 (0.62–3.38) | 0.397 |
| Heart failure | 11 (1.03%) | 15 (1.41%) | 0.38 (-0.56–1.31) | 0.433 | 1.36 (0.63–2.97) | 0.435 |
| Stroke | 4 (0.38%) | 5 (0.47%) | 0.09 (-0.46–0.65) | 0.739 | 1.25 (0.34–4.66) | 0.740 |
| Acute renal failure | 2 (0.19%) | 3 (0.28%) | 0.09 (-0.32–0.51) | 0.655 | 1.50 (0.25–8.98) | 0.657 |
| Urinary tract infection | 45 (4.23%) | 43 (4.04%) | -0.19 (-1.88–1.50) | 0.827 | 0.96 (0.64–1.44) | 0.827 |
| Sepsis | 13 (1.22%) | 28 (2.63%) | **1.41 (0.26–2.56)** | 0.016 | **2.15 (1.13–4.09)** | 0.019 |

RD, risk difference; RR, risk ratio; CI, confidence interval; SD, standard deviations; JPY, Japanese yen. Bold indicates statistically significant.

**Supplementary Table 6.** Outcomes after propensity-score matching for Child-Pugh classes A and B.

|  | Child-Pugh class |  | RD, % (95%CI) | P | RR (95%CI) | P |
| --- | --- | --- | --- | --- | --- | --- |
|  | A | B |  |  |  |  |
| N | 1,012 | 1,012 |  |  |  |  |
| **Outcome** |  |  |  |  |  |  |
| In-hospital death | 15 (1.48%) | 60 (5.93%) | **4.45 (2.79–6.10)** | <0.001 | **4.00 (2.27–7.05)** | <0.001 |
| Length of stay mean, SD | 38.85 (28.07) | 42.62 (32.33) | **3.77 (1.09–6.45)** | 0.006 | **1.10 (1.03–1.17)** | 0.006 |
| Postoperative days mean, SD | 33.12 (25.79) | 36.43 (29.84) | **3.31 (0.84–5.77)** | 0.008 | **1.10 (1.02–1.18)** | 0.008 |
| 30-day readmission | 58 (5.73%) | 82 (8.10%) | **2.37 (0.18–4.56)** | 0.034 | **1.41 (1.02–1.95)** | 0.035 |
| Hospitalization fee mean JPY, SD | 1,939,436 (1,002,526) | 2,104,167 (1,115,397) | **164,730 (73,062–256,399)** | <0.001 | **1.08 (1.04–1.14)** | <0.001 |
| **Complications** |  |  |  |  |  |  |
| **Surgical** |  |  |  |  |  |  |
| Surgical site infection | 2 (0.20%) | 1 (0.10%) | -0.10 (-0.43–0.24) | 0.564 | 0.50 (0.05–5.52) | 0.572 |
| Blood transfusion | 496 (49.01%) | 646 (63.83%) | **14.82 (10.54–19.10)** | <0.001 | **1.30 (1.20–1.41)** | <0.001 |
| **Non-surgical** |  |  |  |  |  |  |
| Acute liver failure | 29 (2.87%) | 71 (7.02%) | **4.15 (2.27–6.03)** | <0.001 | **2.45 (1.60–3.74)** | <0.001 |
| Upper gastrointestinal bleeding | 131 (12.94%) | 121 (11.96%) | -0.99 (-3.82–1.85) | 0.494 | 0.92 (0.74–1.16) | 0.495 |
| Respiratory failure | 37 (3.66%) | 45 (4.45%) | 0.79 (-0.92–2.50) | 0.365 | 1.22 (0.80–1.86) | 0.366 |
| Venous thromboembolism | 42 (4.15%) | 36 (3.56%) | -0.59 (-2.30–1.12) | 0.497 | 0.86 (0.55–1.34) | 0.498 |
| Acute coronary syndrome | 11 (1.09%) | 7 (0.69%) | -0.40 (-1.22–0.43) | 0.346 | 0.64 (0.25–1.64) | 0.350 |
| Heart failure | 15 (1.48%) | 22 (2.17%) | 0.69 (-0.49–1.87) | 0.250 | 1.47 (0.76–2.83) | 0.253 |
| Stroke | 4 (0.40%) | 4 (0.40%) | 0.00 (-0.55–0.55) | 1.000 | 1.00 (0.25–4.00) | 1.000 |
| Acute renal failure | 2 (0.20%) | 6 (0.59%) | 0.40 (-0.15–0.94) | 0.157 | 3.00 (0.61–14.88) | 0.179 |
| Urinary tract infection | 38 (3.75%) | 38 (3.75%) | 0.00 (-1.67–1.67) | 1.000 | 1.00 (0.64–1.56) | 1.000 |
| Sepsis | 27 (2.67%) | 30 (2.96%) | 0.30 (-1.17–1.76) | 0.691 | 1.11 (0.66–1.87) | 0.691 |

RD, risk difference; RR, risk ratio; CI, confidence interval; SD, standard deviations; JPY, Japanese yen. Bold indicates statistically significant.

**Supplementary Table 7.**
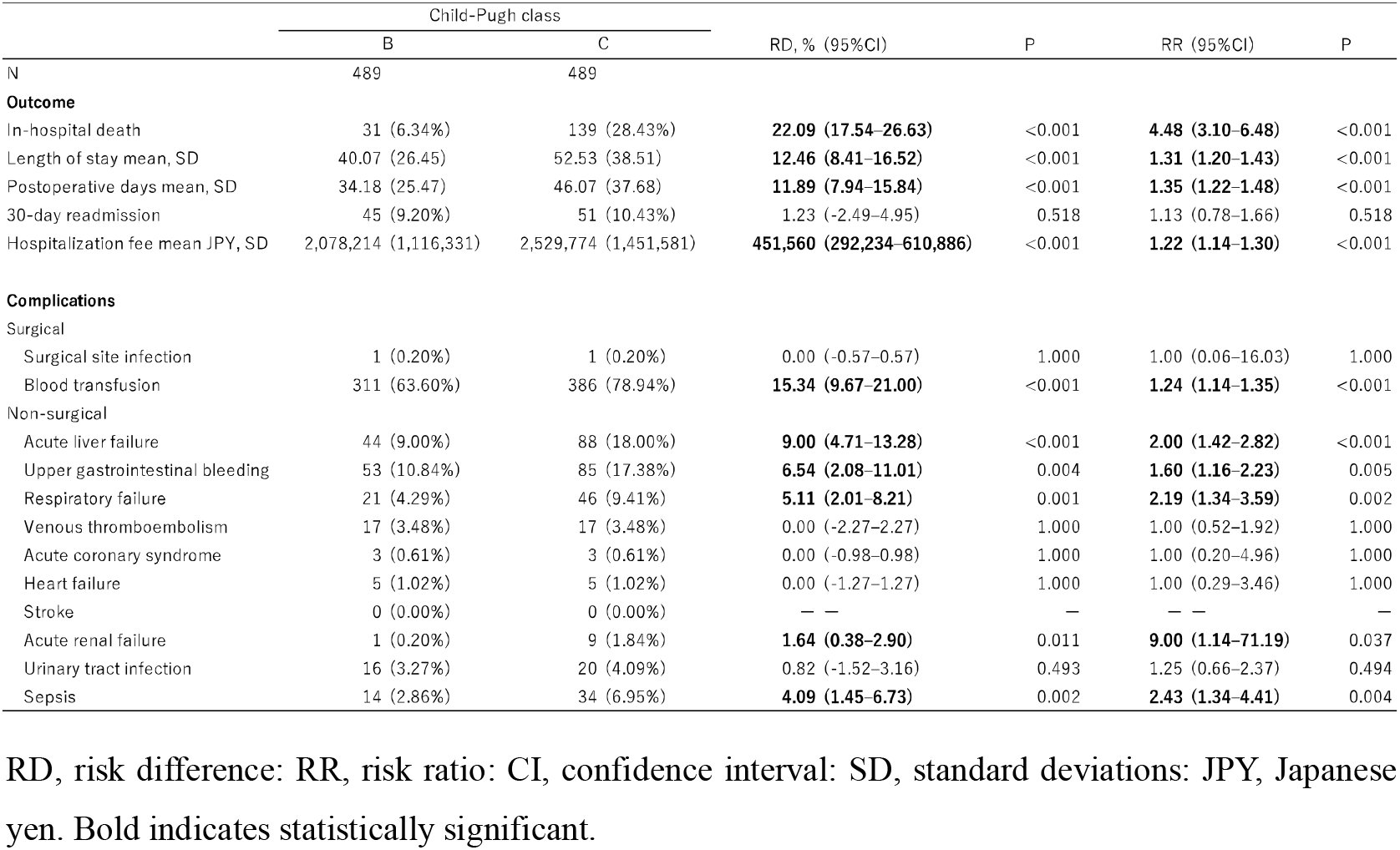
Outcomes after propensity-score matching for Child-Pugh classes B and C.

